# The GLP-1 RA boom: Trends in publicly subsidised and private access in Australia, 2020-2025

**DOI:** 10.1101/2025.10.30.25339120

**Authors:** Michael O Falster, Juliana de Oliveira Costa, Tamara Milder, Joanne Carson, Cade Shadbolt, Brendon L Neuen, Clare Arnott, Andrew Wilson, Min Jun, Nicole Pratt, Sallie-Anne Pearson

## Abstract

**Objectives:** To quantify population-level trends in publicly subsidised and private access of glucagon-like peptide-1 receptor agonists (GLP-1 RA) in Australia.

**Design:** Longitudinal descriptive study using routinely collected data.

**Setting:** National pharmaceutical sales and Pharmaceutical Benefits Scheme (PBS) dispensing claims for GLP-1 RA listed for type 2 diabetes (T2D), May 2020-April 2025.

**Main outcome measures:** We measured GLP-1 RA use according to the total number of units sold/dispensed and defined daily dose (DDD)/1000 population/day. We used sales data to quantify total population use, and PBS dispensings to quantify subsidised access. We estimated private access as the difference between total sales and PBS dispensings.

**Results:** Since May 2020, total sales of GLP-1 RA in Australia increased almost 10-fold, reaching approximately half a million units sold each month in 2024/25, despite significant disruptions to access during shortages. Most growth was in private access, driven initially by semaglutide then rapid uptake of tirzepatide following its market approval. In the year May 2024-April 2025, over 6 million units of GLP-1 RA were sold, predominantly semaglutide (63.3%) and tirzepatide (30.7%); 47.8% of all GLP-1 RA were accessed via the private market, including 26.9% of semaglutide and all tirzepatide. In this period, GLP-1 RA use was estimated to cover 18 out of every 1000 Australians at a standard daily maintenance dose, with 6 out of every 1000 accessing these medicines privately.

**Conclusions:** Australian sales of GLP-1 RAs increased dramatically over the past five years. Half of GLP-1 RA in 2024/25 were accessed in the private market, with an estimated 180,018 to 239,724 Australians accessing GLP-1 RAs privately each month. Our findings underscore the substantial pressures on health systems in meeting the growing demand for these medicines, including rapidly shifting markets, significant demands outside of subsided care, and the need to ensure equitable access and outcomes of use.

**Summary box:** *What is already known on the topic:* Novel GLP-1 RA medicines have significant cardio-renal benefits, including weight loss for people with type 2 diabetes and obesity. There is strong consumer demand for health systems to subsidise these treatments for obesity, yet subsidised access in high-income countries has generally been limited to people with type 2 diabetes. The extent of private market use is largely unknown.

*What this study adds:* Almost half of GLP-1 RA medicines indicated for type 2 diabetes in Australia are accessed on the private market, most likely for weight loss. There were rapidly shifting trends within subsidised and private markets, with disruptions from global shortages and listing of new medicines. Our findings quantify the potential scope of expanding indications and highlight health system challenges for managing equitable use outside of subsidised healthcare.

## Introduction

Glucagon-like peptide-1 receptor agonists (GLP-1 RAs) are reshaping the treatment paradigm for chronic disease.^1^ By acting on multiple metabolic pathways, they help lower blood glucose and reduce risk of cardiovascular and kidney complications.^2, 3^ Originally developed for the treatment of type 2 diabetes (T2D), their benefits now extend to people with obesity, cardiovascular and chronic kidney disease.^2, 4-6^ GLP-1 RA and dual GLP-1/glucose-dependent insulinotropic polypeptide (GIP) receptor agonists have transformed treatment for obesity with clinical trials reporting clinically meaningful weight loss of up to 17.8%.^7, 8^ Access to these medicines has enormous public health benefits, given the interconnected pathophysiology of obesity, T2D, chronic kidney and cardiovascular disease.^1, 9^

While GLP-1 RAs are approved by medicine regulators to treat multiple conditions, subsidised access remains tightly controlled in many health systems due to their considerable budgetary impact.^10-12^ As such, subsidised access does not always reflect evolving consumer demand, particularly for obesity.^7^ In Australia, GLP-1 RAs products have been approved for use in either T2D or obesity, with select products subsidised through the Pharmaceutical Benefits Scheme (PBS) only for people with poorly controlled T2D who meet specific criteria (e.g. prior use of sodium-glucose cotransporter 2 inhibitor [SGLT2i]; use in combination with metformin, sulfonylurea or insulin).^13^ Many high-income countries including Canada, Germany, Netherlands only subsidise GLP-1 RA for T2D, while some jurisdictions including the UK, France and Japan subsidise access for obesity under restricted indications.^11^ Selected US health insurance plans including Veterans Affairs subsidise access to GLP-1 RA medicines for obesity.^11^

International experience points to a sharp rise in consumer demand for these medicines - particularly for weight loss, where access is primarily through the private market. The popularity is driving prescribing outside of the subsidised indications,^14^ shortages in supply,^7, 15, 16^ and disruptions to therapy for people with T2D and obesity alike.^17-21^ These conditions disproportionately affect people from low socio-economic groups, with barriers to access potentially deepening inequities in access to care and health outcomes.^1, 7^ While data on subsidised access to GLP-1 RA is available in many countries, there is a dearth of information on private, non-subsidised use.

In Australia, access to GLP-1 RA for weight loss (without T2D) is only available through private prescriptions, with limited reimbursement in select private health insurance plans.^7^ Some medicines, such as tirzepatide, are not PBS listed and only available through private prescription. Given the cost advantages of accessing GLP-1 RAs for T2D via the PBS (maximum cost AUD$31.60 versus up to $200-$600 per month), it is likely most private access of PBS-listed medicines are for weight loss. Our aim was to quantify trends in use of GLP-1 RAs medicines listed for T2D in Australia, overall and according to publicly subsidised and private market access.

## Methods

### Design and setting

We conducted a longitudinal descriptive study using national data on sales of prescribed GLP-1 RA medicines and Pharmaceutical Benefits Scheme (PBS) dispensing claims to estimate subsidised use through Australia’s universal health insurance scheme, as well as private access outside this scheme, May 2020 – April 2025.

In Australia, the Therapeutic Goods Administration (TGA) is responsible for regulatory (market) approval for medicines, while subsidised access is managed through the PBS. To gain PBS-listing, a medicine must be recommended by the Pharmaceutical Benefits Advisory Committee (PBAC) based on its cost-effectiveness for a specific indication. If a PBS-listed medicine is used outside the PBS-listed indication, prescribers are required to write private prescriptions, requiring patients to pay the full cost.^22^ As of July 2025, PBS-eligible persons pay a maximum co-payment of $31.60 for each PBS item dispensed and concessional beneficiaries (e.g. pensioners, people with disabilities) pay a maximum of $7.70.

### Data access and medicines of interest

#### Sales data

We used data on sales of prescribed medicines provided by IQVIA Inc. (https://www.iqvia.com/). This company collects information on sales of medicines to community pharmacies, hospitals and other settings (e.g., aged-care, medical centres, ambulances, carceral settings) by pharmaceutical wholesalers and manufacturers. These data include the number of packs of products sold nationally each month, including information on formulation, strength, quantity and setting (hospital, community pharmacy) of each sale. IQVIA data ascertains around 97% of the Australian pharmacy and hospital market.^23^

We accessed sales data on all GLP-1 RA listed for use in T2D, including tirzepatide (a dual GLP-1/GIP RA with listing for both T2D and obesity), exenatide, dulaglutide, and specific products for semaglutide and liraglutide listed for T2D. Data were not provided on GLP-1 RA products specifically listed for obesity (semaglutide [Wegovy] and liraglutide [Saxenda]).

#### Pharmaceutical Benefits Scheme (PBS) data

PBS data includes information on PBS-listed medicines dispensed in community pharmacies, private hospitals, and on discharge from public hospitals in most states under PBS-specified indications. PBS data do not capture private prescriptions or medicines supplied to inpatients in public hospitals. We used publicly available data on PBS-subsidised medicine dispensings.^24^ This includes summarised information on monthly dispensings through PBS and Repatriation PBS (hereafter referred to collectively as the PBS). We identified corresponding PBS-listed medicines to those in the sales data. (Supplementary Table 1).

Table 1 details a timeline of TGA approval and PBS-listing for the medicines of interest. Tirzepatide and liraglutide are TGA approved but not PBS listed, and exenatide was removed from the Australian market in 2022. A summary of supply shortages from the Therapeutic Goods Administration (TGA) Medicine Shortage Reports Database and de-listings from PBS is provided in Supplementary Table 2.

**Table 1:** Timeline of GLP-1 RA Therapeutic Goods Administration approval and PBS-listing for T2D in Australia.

| Medicine | TGA approval |  | PBS-listing * |
| --- | --- | --- | --- |
|  | Indication | Date |  |
| Exenatide | T2D | June 2007 | August 2010 |
|  | T2D | December 2012 | September 2016 |
| Liraglutide | T2D | August 2010 | - |
|  | Obesity † | December 2015 | - |
| Dulaglutide | T2D | January 2015 | June 2018 |
| Semaglutide | T2D | August 2019 | July 2020 |
|  | Obesity † | August 2024 | - |
| Tirzepatide | T2D | December 2022 | - |
|  | Obesity | September 2024 | - |

\* Listed on the Australian Pharmaceutical Benefits Scheme with restrictions for use in patients with poorly controlled T2D who are currently receiving metformin, insulin and/or sulfonylurea but not receiving subsidised SGLT2i class medications.
† Sales data on products specifically indicated for obesity (Wegovy, Saxenda) were not provided for this study as they are not indicated for T2D
Abbreviations: PBS, Pharmaceutical Benefits Scheme; T2D, type 2 diabetes; TGA, Therapeutic Goods Administration

### Measuring GLP-1 RA use

We measured medicine use two ways. First, we used the total number of packs/units sold and dispensed. This represents the base unit of measurement in both the IQVIA sales and PBS claims data respectively, with one pack/unit typically provided when filling a single prescription. Packs contain differing strengths and quantities.

Second, we used Define Daily Dose, a statistical unit by the World Health Organization (WHO) to standardise drug consumption across products with varying formulations, volumes and potencies. The DDD represents the assumed average maintenance dose per day for a medicine used in its main indication in adults, allowing for comparisons across medicines and populations. For each product, we calculated the total milligrams of active ingredient per unit (e.g. per pen or vial) based on formulation, strength, and pack size. We then derived total DDDs per month by multiplying the strength (mg) by the number of units dispensed, and dividing by the WHO-defined DDD for that product (Supplementary Table 1). For these medicines the DDD was based on an indication for T2D, except tirzepatide which is based on dual indication for T2D and obesity. To quantify population-standardised use, we calculated DDDs per 1,000 population per day (DDD/1000/day), which represents the average number of people per 1,000 population receiving a standard daily dose for T2D. We sourced annual population from the Australian Bureau of Statistics.

### Statistical analysis

We used IQVIA data to quantify total sales of GLP-1 RA medicines. Where total sales for a medicine had a negative value (e.g. due to returns, inter-store transfers) we quantified these as zero. We used PBS dispensings to quantify subsidised access. We estimated private access by subtracting the total number of PBS dispensings from total units sold. While there may be a lag between sales to pharmacies and subsequent dispensing, we assume this gap will be minimal given the supply shortages and likelihood stock will be depleted. In some instances, we observed the number of monthly PBS dispensings were larger than the total number of monthly sales (e.g. when exenatide was removed from the market and when there were large fluctuations in semaglutide supply). We assumed this was depletion of stock and counted private access as zero. At these times the total estimated use (PBS subsidised + private access) is higher than the total sales, however these differences were generally minor (Supplementary Figures 1-2).

We described monthly trends in use of GLP-1 RA across May 2000 – April 2025, as both total sales as well as both PBS-subsidised and private access. We also measured use in the most recent year of data available (May 2024 – April 2025).

### Patient and public involvement

We held a forum on June 5 2025 with a panel of healthcare consumers, facilitated through Health Consumers New South Wales, to discuss experiences accessing cardiometabolic medicines including GLP-1 RA. The panel included people with conditions including obesity, T2D, cardiovascular and kidney disease. Consumers flagged the importance of investigating differences in subsidised and private use, as well as potential implications on access and equity across Australia. At the completion of the study, members of the consumer panel were invited to comment on our research findings. We have also co-developed a lay summary of our findings that will be disseminated to the consumer panel and the broader community at the time the research is published.

## Results

### Trends in GLP-1 RA sales

Total GLP-1 RA sales increased almost 10-fold from 57,941 units in May 2020 to 496,875 units in April 2025 (Figure 1). Within this period, there was a gradual increase in sales up to June 2022 (282,002 units), a rapid decrease during times of shortages (down to 59,315 units in January 2023) and a subsequent sharp rise (up to 516,156 units in May 2023). Total sales fluctuated across the following months, but with a slight increasing trend in 2024.

**Figure 1:**
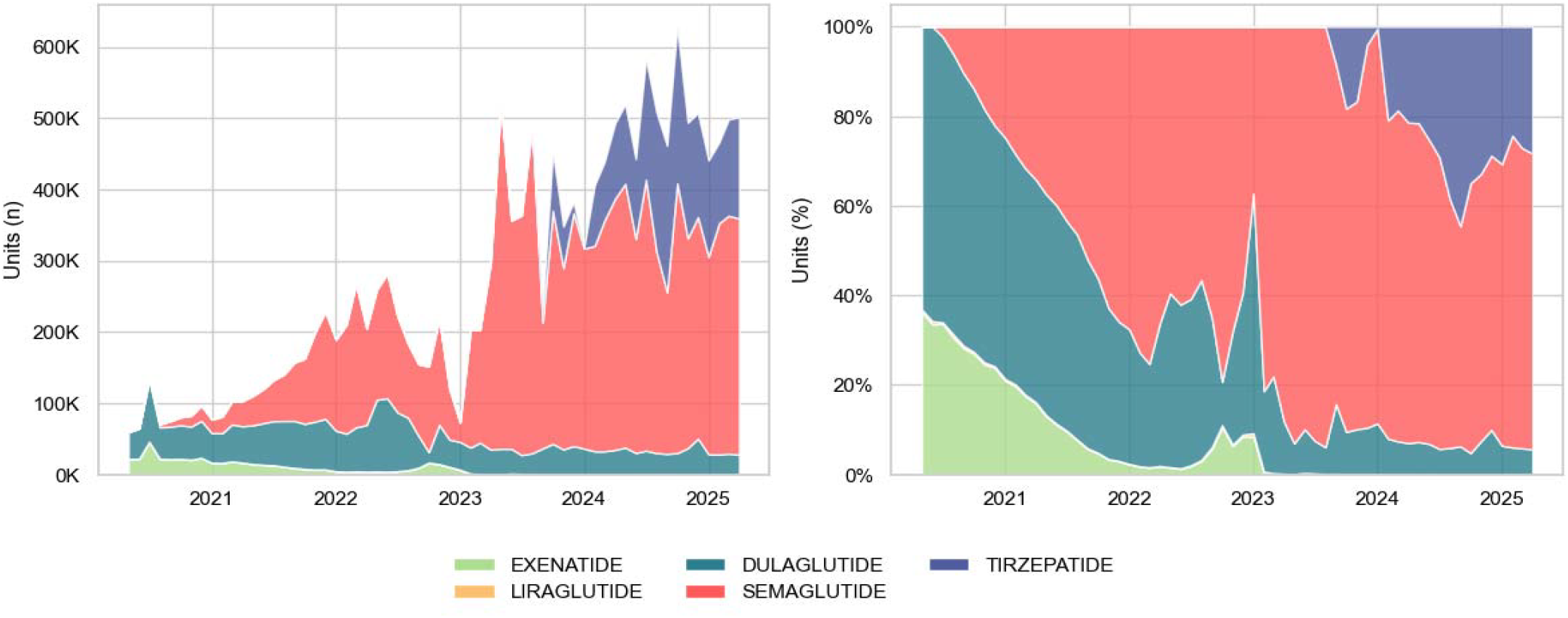
Monthly trends in the number of GLP-1 RA medicine sales in Australia, May 2020 – April 2025, according to medicine type. (a) Number of units. (b) Proportion of total units Data Source: IQVIA Solutions Australia and PBS dispensing claims

The increasing trends were driven primarily by an increase in semgalutide (Figure 1). Following market listing in early 2020, sales of semaglutide increased to 175,095 units in June 2022, dropped during shortages to 15,455 units in January 2023, followed by a sharp increase (to 484,419 units in May 2023) with fluctuating monthly sales after that time. There was a rapid increase in sales of tirzepatide following market introduction in 2024, which peaked at 219,701 units in October 2024. From mid-2023 onwards these semglutide and tirzepatide combined comprised over 90% of total GLP-1 RA sales. Sales of dulaglutide increased gradually from May 2020 (37,536 units) to June 2022 (103,143 units), after which sales decreased to relatively sustained use from 2023 onwards (between 20,000-50,000 units a month). Liraglutide sales started at 364 units in May 2020, remaining low across the period. Exenatide sales started at 20,041 units in May 2020, gradually decreasing before sales stop in mid-2023.

### Trends in PBS-subsidised and private access

PBS-subsidised access increased relatively steadily over the 5-year period (Figure 2), from 58,902 units in May 2020 to 279,118 units in April 2025. We observed a drop in PBS-subsidised access during the period of sales shortages (down to 69,379 units in January 2023), from May 2023 onwards access fluctuated slightly between 200,000-300,000 units a month, with a slight increasing trend over time. The predominant PBS-subsidised medicine from 2023 onwards was semaglutide (Figure 3, Supplementary Figure 2).

**Figure 2:**
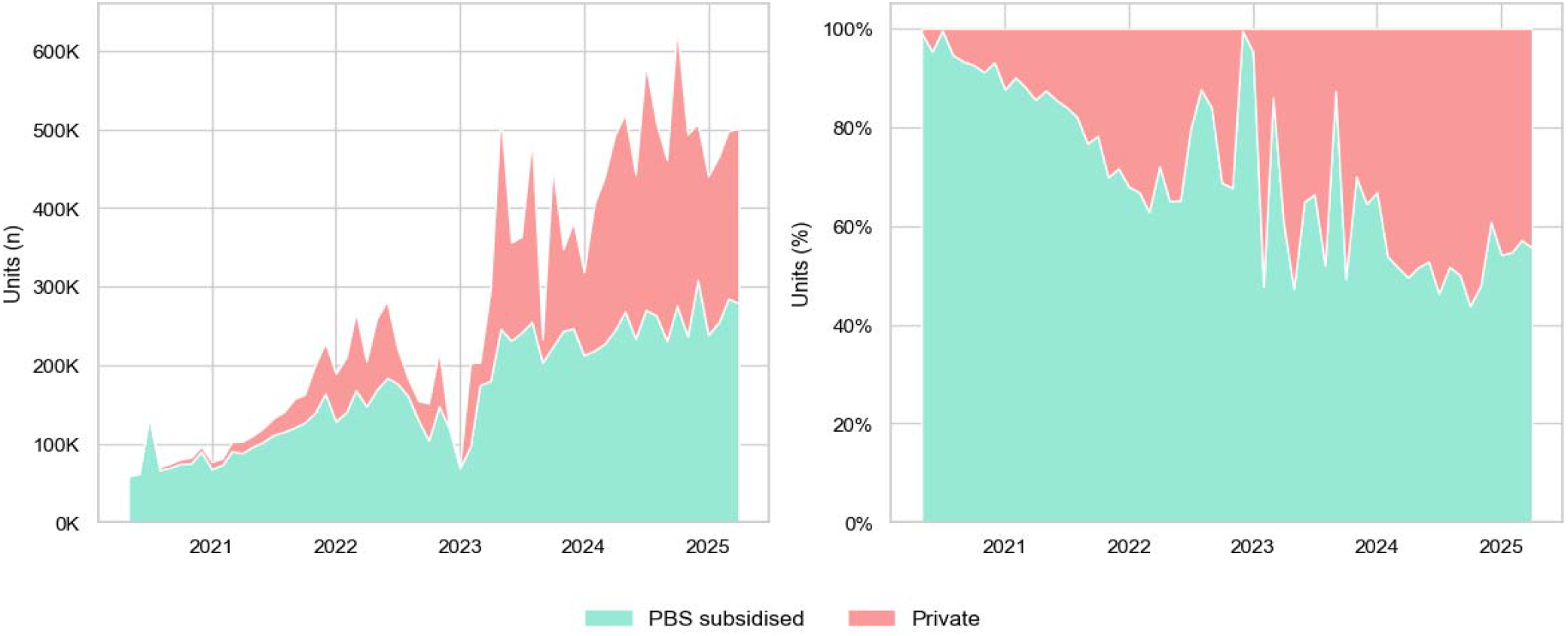
Monthly trends in PBS-subsidised and private access to GLP-1 RA medicines in Australia, May 2020 – April 2025. (a) Number of units; (b) Proportion of total estimated access. Data Source: IQVIA Solutions Australia and PBS dispensing claims

**Figure 3:**
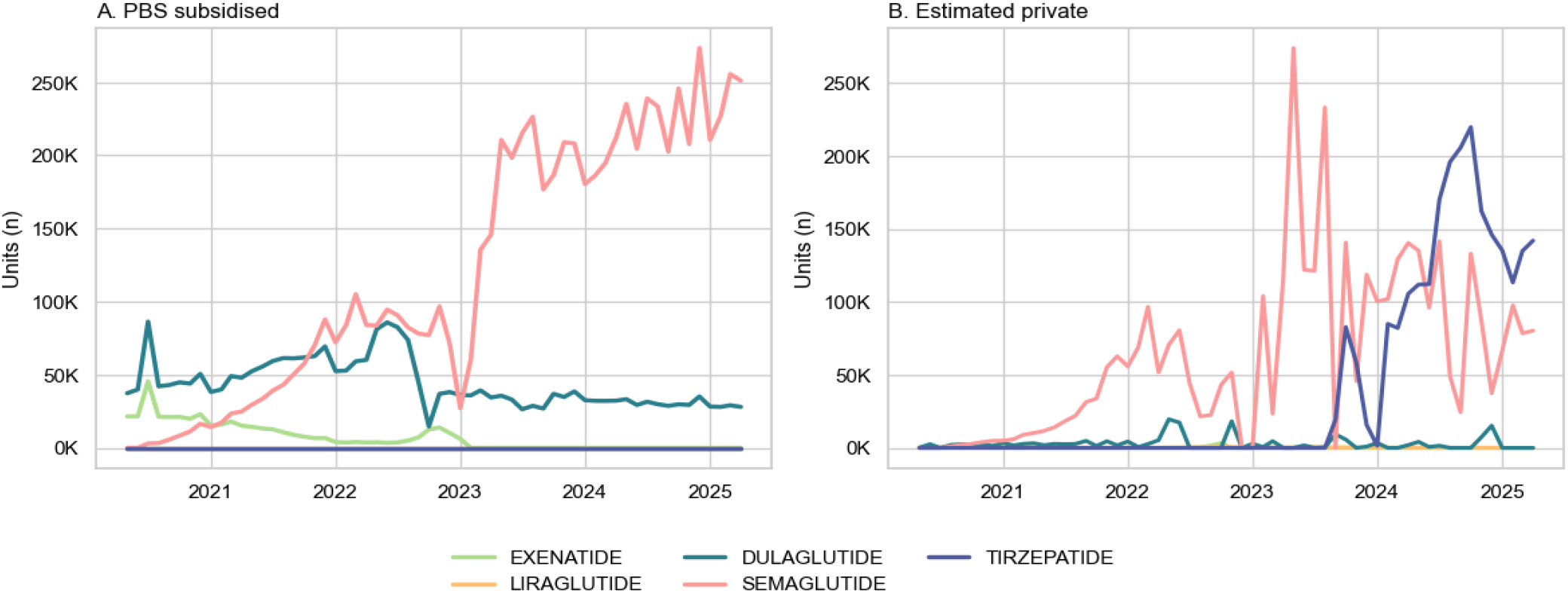
Monthly trends in GLP-1 RA medicines accessed in Australia, May 2020 – April 2025. (a) PBS-subsidised access; (b) Estimated private access. Data Source: IQVIA Solutions Australia and PBS dispensing claims

Estimated private access increased from no use in May 2020 to 43.8% of total estimated use in April 2025 (Figure 2). During this time, private access gradually increased through late 2021 and mid-2022, hitting 34.9% of total estimated use in May 2022. At the lowest period of total supply (December 2022 -January 2023) there were more PBS dispensings than sales (Supplementary Figure 1), resulting in no estimated private access. There were large monthly fluctuations in estimated private supply from mid-2023 onwards, mostly reflecting fluctuations in total sales during this period. Up to mid-2023 the predominant medicine among privately accessed GLP-1 RA was semaglutide (Figure 3, Supplementary Figure 2). Private access to semaglutide decreased after tirzepatide entered the market, with tirzepatide becoming the predominant medicine from June 2024 onwards.

We found similar patterns in use over time according to DDDs/1000/population/day (Supplementary Figures 4-6).

### GLP-1 RA access in Australia, May 2024 – April 2025

In the period May 2024 to April 2025 there was an estimated 6,022,882 GLP-1 RA units accessed in Australia (Table 2). Of these, 63.3% were semaglutide, 30.7% were tirzepatide, 6.0% dulaglutide, and <1% liraglutide. Of all estimated GLP-1 RA access, 52.2% were PBS-subsidised and 47.8% were accessed privately. The proportion of private-market access differed across medicines: with 26.9% of semaglutide, all of tirzepatide and liraglutide, and none of dulaglutide (where PBS dispensings were greater than the number of sales).

**Table 2:** Total estimated access to GLP-1 RA medicines in Australia May 2024-April 2025, and estimated use according to PBS-subsidised and private access.

|  | Total estimated<br>access * | PBS-subsidised |  | Private |  |
| --- | --- | --- | --- | --- | --- |
|  |  | n | % of<br>total | N | % of total |
| Units/Packs |  |  |  |  |  |
| All GLP-1 RA | 6,022,882 | 3,146,179 | 52.0 | 2,876,703 | 47.8 |
| Semaglutide | 3,812,004 | 2,785,760 | 73.1 | 1,026,244 | 26.9 |
| Tirzepatide | 1,848,841 | - | - | 1,848,841 | 100 |
| Dulaglutide | 360,419 | 360,419 | 100 | 0 | 0 |
| Liraglutide | 1,618 | - | - | 1,618 | 100 |
| DDD/1000/day |  |  |  |  |  |
| All GLP-1 RA | 17.62 | 11.05 | 62.7 | 6.57 | 29.8 |
| Semaglutide | 12.90 | 9.59 | 74.3 | 3.31 | 25.7 |
| Tirzepatide | 3.25 | - | - | 3.25 | 100 |
| Dulaglutide | 1.46 | 1.46 | 100 | 0.00 | 0 |
| Liraglutide | <0.01 | - | - | <0.01 | 100 |

\* As the total of PBS subsidised and private access
Data Source: IQVIA Solutions Australia and PBS dispensing claims

Overall GLP-1 RA use was 17.62 DDD/1000/day– indicating that on average, GLP-1 RA was estimated to cover 1.8% of the total Australian population at a standard daily maintenance dose. We observed a similar pattern of PBS-subsidised and privately accessed DDDs to units sold (Table 2), although private market access comprised a lower proportion of DDDs for total GLP-1 RA use (29.8%) than for units sold. This is because the majority of tirzepatide access was at lower strengths, typically used as a starting dose. Private market access of GLP-1 RA was 6.6 DDD/1000/day, estimated to cover 0.7% of the Australian population at a standard daily maintenance dose.

## Discussion

We observed a rapid shift in GLP-1 RA use in Australia, with sales increasing almost tenfold from early 2020 to early 2025. This growth was primarily rapid adoption of new therapies semaglutide and tirzepatide, with significant disruptions in supply over time. By 2024/25 nearly half of GLP-1 RA access was on the private market - likely driven by demand for use in weight loss, given the cost advantages of PBS-subsidised access for T2D. Our findings underscore the urgent need for health systems to respond to the rising demand for these medicines – including challenges such as expanding reimbursement beyond T2D, promoting equitable access, and strengthening surveillance of care and outcomes, particularly where use is off-label and/or through the private market use.

The key strength of our study is its whole-of-system perspective, including national data on GLP-1 RA use both within and outside of publicly subsidised care. It is the first study to quantify the extent of both subsidised and private use, the latter having little visibility given only subsidised use is typically captured in routine surveillance. There is no doubt, however, we have underestimated GLP-1 RA use in the context of weight loss, given our sales data only included medicines labelled for T2D. The use of semaglutide for obesity likely represents an addition to the overall market demand, given we found no corresponding reduction in overall use at the time Wegovy was introduced to the Australian market in August 2024. Our estimates should be considered the minimum estimated GLP-1 RA use in Australia. There is limited data on private market use of GLP-1 RA medicines for which we can compare our results, however our findings are consistent with international studies showing increasing use of GLP-1 RA outside T2D; for example 20% of medicines dispensed in a US health insurer being indicated for obesity.^25^ Recent studies are demonstrating high levels of off-label use, such as one third of people initiating semaglutide labelled for T2D in Denmark having no history of T2D,^14^ and over 90% of Australian women of child-bearing age initiating GLP-1 RA in general practice in 2022 having no evidence of T2D.^26^ It is possible a large proportion of PBS-subsidised access in our study may also be among people without T2D.

Our study also contains highly contemporary data, presenting novel insights on recent disruptive events such as periods of GLP-1 RA shortages and the recent listing of tirzepatide. Our trends are broadly consistent with global studies demonstrating rapidly increasing use of GLP-1 RA,^14, 25, 27-30^ particularly emerging demand for semaglutide.^28, 31^ Very few studies have more recent data (2023 on), primarily from US insurance companies. ^25, 29^ These also show a sharp increase in tirzepatide dispensing following market entry in both people with and without T2D.^25, 29^ While supply shortages in GLP-1 RA use are well known, few studies have shown their impact on population-level use. We found a sharp drop – almost 80% - in use in Australia from mid-late 2022. These shortages impacted both PBS-subsidised and private access, with PBS dispensings outnumbering sales at the peak of these shortages. More modest drops were seen in the US,^32^ with very little impact in some health insurance schemes.^25^ These discrepancies highlight the vulnerabilities of different markets during times of supply chain shortages.

Key limitations of our study are that we did not have data on GLP-1 RA products labelled for obesity, or patient-level data to determine characteristics of people using these medicines (e.g. age, sex, socio-demographic factors) or the indication for prescribing (e.g. off-label use in people without T2D, use outside regulatory approvals among people with neither T2D or obesity).

### Implications for health policy and patient care

Reimbursement of GLP-1 RA for obesity remains a challenge worldwide.^12^ Most high-income countries do not currently subsidise GLP-1 RA for this indication, with concerns about the budget impacts and cost-effectiveness of these medicines^10, 11^ – particularly given the high prevalence of the condition (one-third of Australian adults have obesity)^33^ and need for long-term therapy to maintain weight loss. In the US, it is estimated 93 million people may be eligible for GLP-1 RA for obesity.^34^ Despite this, GLP-1 RA for T2D are already one of the costliest (in terms of total budget impact) medicines worldwide. For example, in the US and Denmark semaglutide for T2D is already the top medicine in terms of health care expenditure;^31, 35^ while in Australia it is ranked 11^th^ in terms of PBS expenditure.^36^ Both Australia and Denmark have recently enacted tightened restrictions to contain costs.^11, 13^

Given most GLP-1 RA products in this study were not TGA approved for obesity (with the exception of tirzepatide), their use for weight loss (without T2D) is considered off-label. While semaglutide is TGA approved for obesity and the clinical benefits have been established,^5^ the large private access in this study is for the products specifically listed for T2D. Obesity is a global pandemic, closely linked to the development of atherosclerotic disease, heart failure, T2D and many other chronic health conditions. There is an urgent need of effective strategies to address obesity, and very few safe and effective pharmacotherapies to treat this highly morbid condition.^7, 9^

Subsidy decisions in Australia are contingent on demonstrable cost-effectiveness. A key factor in for submissions is to estimating the budgetary impact of subsidising these medicines, including the size of the eligible treatment population; this was a key driver of uncertainty in the recent recommendation not to recommend the PBS subsidy of Wegovy.^37^ Using the figures generated in our study, we estimate that between 482,788 and 501,907 Australians are using GLP-1 RA each month. The lower estimate is based on DDDs, where we found access to GLP-1 RA medicines equivalent to 1.8% (17.62/1000 pop/day) of Australians per day at a standard maintenance dose. The upper estimate is based on the number of medicines dispensed and the assumption people fill on average one dispensing a month. Similarly, we estimate that between 180,018 and 239,725 Australians are purchasing GLP1-RA privately each month, likely for weight loss. Given our study does not include Wegovy, and that some subsidised access may potentially be outside of PBS restrictions for T2D^14^, our estimates should be considered the minimum current market for weight loss outside of T2D.

Our findings raise equity concerns. Overweight and obesity are major contributors to morbidity and mortality, are more prevalent among First Nations people, as well as people living in regional areas and socioeconomically disadvantaged groups.^33^ Like people with T2D, these populations face greater health disparities and access barriers to preventive care. The lack of subsidised access to GLP-1 RA for weight loss risks entrenching existing health inequities. Socioeconomic disparities in prescribing these medicines for obesity and T2D are already documented in Australia and the US. ^32, 38, 39^ Given the higher out-of-pocket costs of private access, further gaps in use and adherence to treatment in the private market are likely.

Our study raises questions about the how the health system should respond to market-driven demands for medicines with unsubsidised indications. Given data on private market dispensings are not included in routine data collections (e.g. PBS claims), people using these medicines, and their outcomes, are not visible to polcymakers.^7^ This limits capacity for surveillance, monitoring uptake, best-practice use, and real-world evidence on safety and effectiveness. There is a need to ensure people using these medicines receive appropriate counselling, for example about lifelong use to maintain weight loss, potential risks associated with discontinuation (e.g. muscle and bone loss^40^), and unknown long-term safety of these medicines, which need to be communicated with prescribers and patients alike.

### Future research

Our study has demonstrated rapidly shifting use of GLP-1 RA medicines listed for T2D, and that almost half of contemporary access is within the private market, most likely for weight loss. However, multiple challenges remain. Given the size of the private market, it is likely much of the current research and reporting on uptake of GLP-1 RA in Australia and internationally significantly under-estimates utilisation. We need greater capacity to measure private market use, such as prescribing in primary and specialist care, as well as dispensing of private prescriptions. Given most high-income countries do not currently subsidise GLP-1 RA use for obesity, international comparisons are required to understand the true extent of the private market. With ever increasing demand for these medicines, we need ongoing surveillance to understand the impact of shifting markets, ensure equity of access, and the impacts on patient adherence and long-term outcomes.

## Data Availability

Sales data were provided by IQVIA Solutions Australia. Access to these data by other individuals are not permitted without permission of the data custodians. PBS dispensing data are available online.

https://www.pbs.gov.au/info/statistics/dos-and-dop/dos-and-dop

## Acknowledgements

This research is supported by the National Health and Medical Research Council (NHMRC) Ideas Grants (grant numbers 2002889, 1183273) and National Health and Medical Research Council (NHMRC) Medicines Intelligence Centre of Research Excellence (grant number 1196900). MOF is supported by a National Heart Foundation of Australia Future Leader Fellowship (grant number 105609). BLN is supported by grants from the National Health and Medical Research Council of Australia, Medical Research Future Fund, Ramaciotti Foundation, and New South Wales Health.

We acknowledge Melisa Litchfield for her assistance with project data and governance. We acknowledge Scott Thrift and Pam Davis for their insights as Consumers into the study question and interpretation of results. The statements, findings, conclusions, views, and opinions contained and expressed in this publication are based in part on data obtained under license from IQVIA Solutions Australia, National Sales Data, May 2020 – April 2025, All Rights Reserved. The statements, findings, conclusions, views, and opinions contained and expressed herein are not necessarily those of IQVIA Solutions Australia, or any of its affiliated or subsidiary entities.

