## Supplementary Material for "The GLP-1 RA boom: Trends in publicly subsidised and private access in Australia, 2020-2025"

### Supplementary Table 1. GLP-1 RA available in Australia

| Medicine | Indication | Administration schedule | Maintenance dose range * | WHO DDD | PBS item codes |
|---|---|---|---|---|---|
| Exenatide | T2D | 10 - 20 μg twice daily | 0.01 - 0.02 mg | 0.015 mg | 03423E, 03424F |
| | T2D | 2 mg weekly | 0.286 mg | 0.286 mg | 10888C |
| Liraglutide | T2D | 1.8 mg daily | 1.2 - 1.8 mg | 1.5 mg | Not subsidised |
| | Obesity † | 3 mg daily | 3 mg | 1.5 mg | Not subsidised |
| Dulaglutide | T2D | 0.75 - 4.5 mg weekly | 0.11 - 0.64 mg | 0.16 mg | 11364D, 14150R |
| Semaglutide | T2D | 0.5 - 2 mg weekly | 0.07 - 0.28 mg | 0.11 mg | 12080T, 14149Q, 14844G, 14846J, 12075M, 14163K |
| | Obesity † | 1.7 - 2.4 mg weekly | 0.24 - 0.34 mg | - | Not subsidised |
| Tirzepatide | T2D<br>Obesity | 5 - 15 mg weekly | 0.7 - 2.14 mg | 1.4 mg<br>- | Not subsidised<br>Not subsidised |

* Maintenance dose range obtained from product information produced by pharmaceutical companies and/or regulatory agencies.
† Sales data on medicines specifically indicated for obesity (Wegovy and Saxenda) were not provided for this study.
**Abbreviations:** ATC, Anatomical Therapeutic Chemical (ATC); DDD, defined daily dose; PBS, Pharmaceutical Benefits Scheme; TGA, Therapeutic Goods Administration; WHO, World Health Organisation

Supplementary Table 2. Summary of National GLP-1 RA supply shortages

| ARTG ID | Active drug | Start date | End date | Availability | Reason | Management |
|---|---|---|---|---|---|---|
| 308324 | Semaglutide 1.34 mg/mL | - | - | Reduction in supply until exhausted | Discontinued - Commercial changes/viability | Product to be deleted from 01/12/2025 |
| 315107 | Semaglutide 1.34 mg/mL | 15/04/2022 | 18/07/2025 | Limited Availability. | Unexpected increase in consumer demand. | Shortage resolved from 19/07/2025. Unregistered product approved for supply under Section 19A |
| 446290 | Semaglutide .68 mg/mL | 1/06/2025 | 18/07/2025 | Limited Availability | Unexpected increase in consumer demand | Shortage resolved from 19/07/2025. Unregistered product approved for supply under Section 19A |
| 217965 | Dulaglutide 1.5 mg | 27/06/2022 | 30/06/2026 | Limited Availability. | Manufacturing | Sponsor monitoring stock levels and releasing to wholesalers as equitably as possible. Unregistered product approved for supply under Section 19A |
| 153980 | Liraglutide 6 mg/mL | - | - | Unavailable | Discontinued Commercial changes/viability | Victoza brand drug deleted from 01/12/2024 |
| 407050 | Tirzepatide 5 mg | 22/11/2023 | 31/08/2025 | Unavailable | Commercial changes/viability | Vial form of drug deleted from 13/06/2025. Replaced by multi-dose pen device. |
| 407051 | Tirzepatide 7.5 mg | 22/11/2023 | 31/08/2025 | Unavailable | Commercial changes/viability | Vial form of drug deleted from 13/06/2025. Replaced by multi-dose pen device. |
| 407052 | Tirzepatide 12.5 mg | 22/11/2023 | 31/08/2025 | Unavailable | Commercial changes/viability | Vial form of drug deleted from 13/06/2025. Replaced by multi-dose pen device. |
| 407053 | Tirzepatide 10 mg | 22/11/2023 | 31/08/2025 | Unavailable | Commercial changes/viability | Vial form of drug deleted from 13/06/2025. Replaced by multi-dose pen device. |
| 407054 | Tirzepatide 15 mg | 22/11/2023 | 31/08/2025 | Unavailable | Commercial changes/viability | Vial form of drug deleted from 13/06/2025. Replaced by multi-dose pen device. |
| 407055 | Tirzepatide 2.5 mg | 22/11/2023 | 31/08/2025 | Unavailable | Commercial changes/viability | Vial form of drug deleted from 13/06/2025. Replaced by multi-dose pen device. |

**Abbreviations:** ARTG, Australian Register of Therapeutic Goods; ID, identifier

**Source**: Therapeutic Goods Administration (TGA) Medicine Shortage Reports Database. A national shortage is defined by the TGA as a situation in which the supply of a registered prescription medicine is not expected to meet normal or projected consumer demand in Australia at any time within the next six months. Data on shortages accessed August 2025 at https://www.tga.gov.au/safety/shortages

**Note:** Section 19A of the Therapeutic Goods Act 1989 allows medicines not currently included in the Australian Register of Therapeutic Goods (ARTG) to be imported into Australia and supplied in place of a registered medicine that is unavailable or in short supply or if relevant registered medicines do not exist but an application for a new medicine is being evaluated by the TGA.

Supplementary Figure 1: Monthly trends in total GLP-1 RA sales, PBS-subsidised dispensing, and estimated private access for (a) all GLP-1 RA medicines; (b) semaglutide; (c) dulaglutide; and (d) exenatide. Negative values for private access indicate PBS dispensings exceed sales; estimated private market is measured as zero.

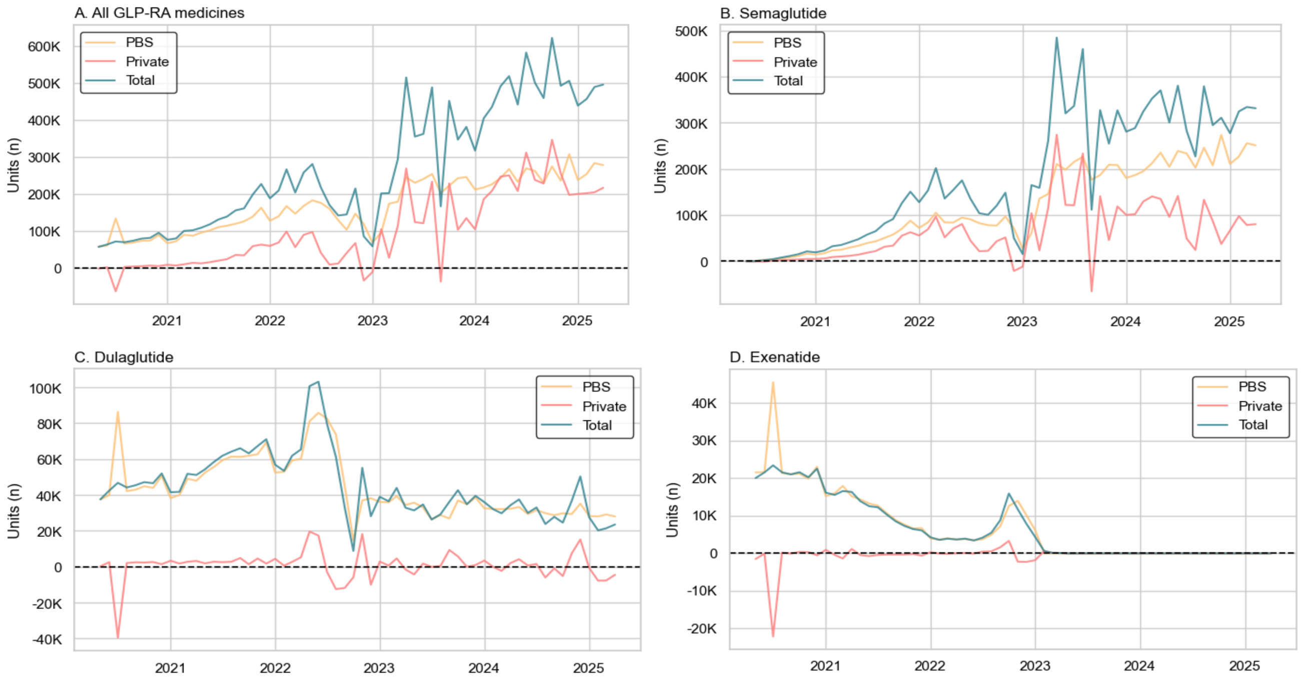


**Data Source:** IQVIA Solutions Australia and PBS dispensing claims

Supplementary Figure 2: Concordance in monthly estimates of GLP-1 RA use when measured as either total GLP-1 RA sales or estimated total use (PBS-subsidised + private access). (a) all GLP-1 RA medicines; (b) semaglutide; (c) dulaglutide; and (d) exenatide. Discrepancies are when PBS dispensings exceed number of sales.

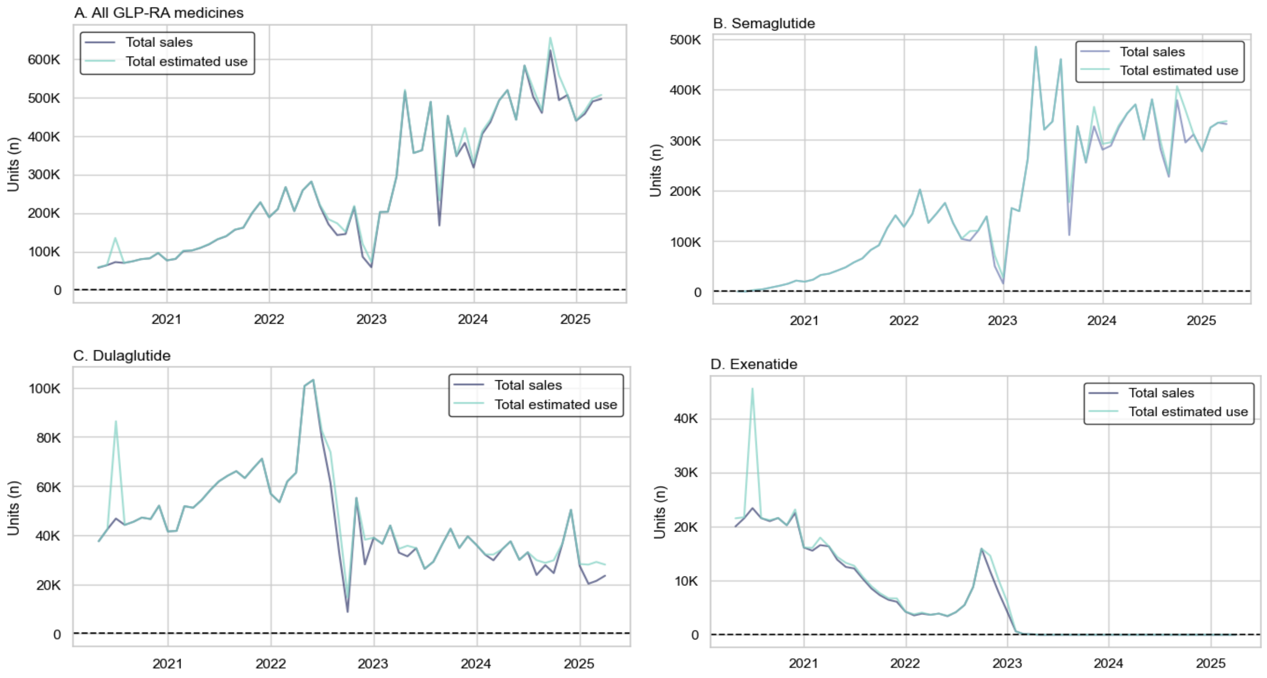


**Data Source:** IQVIA Solutions Australia and PBS dispensing claims

Supplementary Figure 3: Monthly trends in the total of number and proportion of GLP-1 RA medicines used in Australia, according to medicine type and PBS-subsidised and private access, May 2020 – April 2025

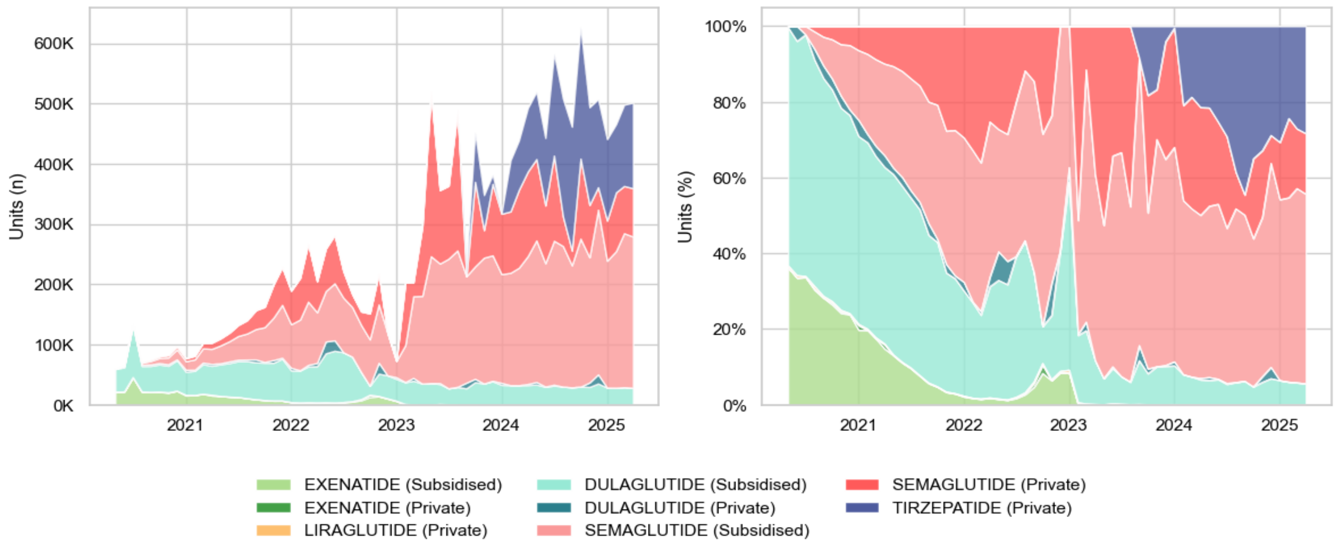


**Data Source**: IQVIA Solutions Australia and PBS dispensing claims

Supplementary Figure 4: Monthly trends in the total Defined Daily Dose (DDD) per 1000 people per day of GLP-1 RA medicine use in Australia, according to PBS-subsidised and private access, May 2020 – April 2025

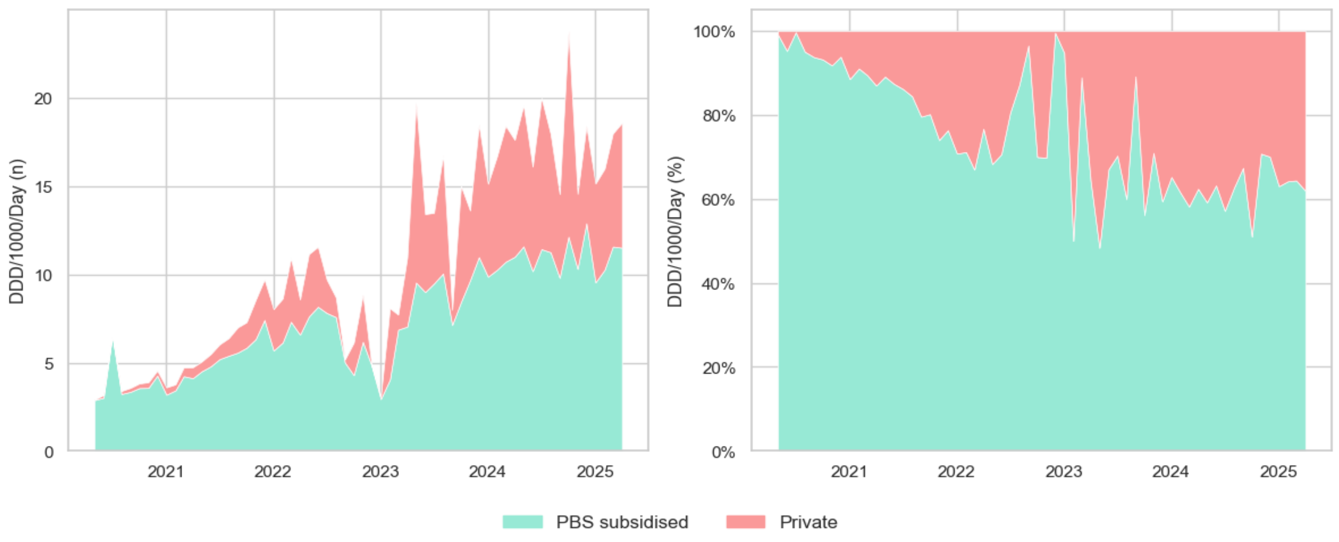


**Data Source:** IQVIA Solutions Australia and PBS dispensing claims

Supplementary Figure 5: Monthly trends in the relative size of Defined Daily dose (DDD) per 1,000 population of PBS-subsidised and privately accessed GLP-1 RA medicines in Australia, May 2020 – April 2025

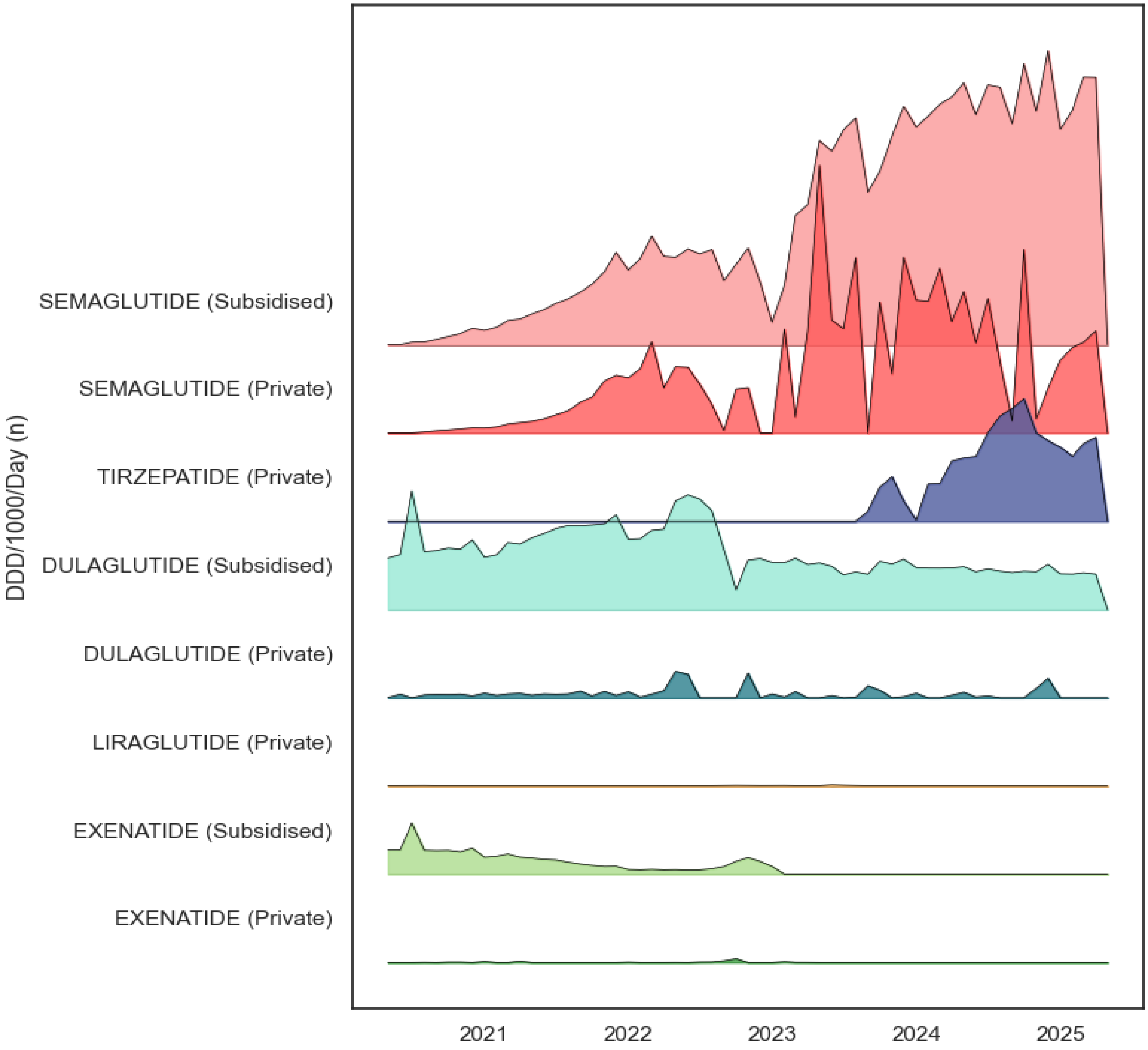


**Data Source:** IQVIA Solutions Australia and PBS dispensing claims

Supplementary Figure 6: Monthly trends in the total Defined Daily Dose (DDD) per 1000 people per day of GLP-1 RA medicine use in Australia, according to medicine type and PBS-subsidised and private access, May 2020 – April 2025

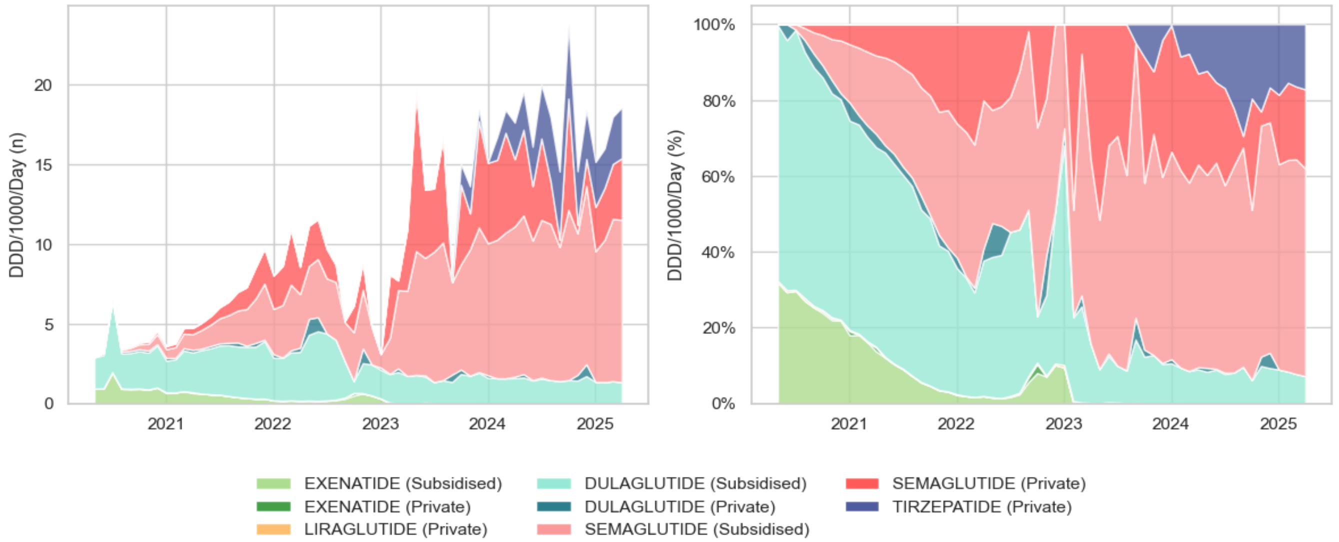


**Data Source:** IQVIA Solutions Australia and PBS dispensing claims
